# Risk Factors Associated with Acute Pancreatitis in Diabetic Ketoacidosis Patients: a 11-year experience in a single tertiary medical center and comprehensive literature review

**DOI:** 10.1101/2025.02.05.25321763

**Authors:** Yifei Chen, Yan Bo, Zhuanzhuan Han, Mengjie Chen

**Affiliations:** The Department of Emergency Medicine, The Affiliated Hospital of Yangzhou University, Yangzhou 225012, Jiangsu Province, China; The Department of Medicine, Northwest Minzu University, Lanzhou 730000, Gansu Province, China; The Department of Medicine, Dalian Medical University, Dalian 116044, Liaoning Province, China

**Keywords:** Diabetic Ketoacidosis, Acute Pancreatitis, Pancreatitis, Risk Factors, Diagnosis

## Abstract

**Objective:** To identify independent risk factors and predictive markers for acute pancreatitis (AP) in patients with diabetic ketoacidosis (DKA).

**Methods:** Here, we reported a 11 year experience in a single tertiary medical center and conduct a systematic review of acute pancreatitis in diabetic ketoacidosis patients cooccurence of DKA and AP. 1941 cases were included. First retrospective cohort study analyzed clinical data from 45 patients with cooccurence of DKA and AP and 45 matched controls with DKA alone admitted to Yangzhou University Affiliated Hospital. Baseline characteristics included BMI, hyperlipidemia history, and serological profiles. Secondly we retrieved clinical studies of cooccurence of DKA and AP from PubMed database system and analysed these clinical data in depth.

**Results:** Significant differences were observed in BMI (26.30 vs. 23.20 kg/m^2^, p = 0.031), hyperlipidemia history (26.7% vs. 4.4%, p = 0.004), abdominal pain duration (3 vs. 0 days, p < 0.001), and lipid profiles [(TC: 7.71 vs. 4.86 mmol/L); (TG: 11.97 vs. 1.60 mmol/L, p < 0.001)]. Logistic regression identified TC [(OR = 1.455, 95% CI(1.196, 1.769)], TG [(OR = 2.046, 95% CI(1.202, 3.484)], and abdominal pain duration [(OR = 3.892, 95% CI(2.173, 6.972)] as independent risk factors. Receiver operating characteristic curve (ROC) analysis demonstrated strong predictive performance (combined AUC = 0.933, sensitivity = 93.3%, specificity = 73.3%). Moreover, we found 11 clinical studies investigating cooccurence of DKA and AP. Cumulatively, 1851 patients were studied, including 3 interventional studies, 1 genetic observational study, and 7 cohort studies exploring risk factors.

**Conclusion:** Elevated TC, TG, and prolonged abdominal pain duration are key predictors of cooccurence of DKA and AP. These factors enhance early diagnosis and clinical management in Yangzhou, China.

## 1 Introduction

The current clinical evidence supporting the existence of a distinct set of risk factors for the co-occurence of diabetic ketoacidosis (DKA) and acute pancreatitis (AP), called DKA-AP, is inconclusive. The interplay between DKA and AP disease makes the clinical diagnosis of the DKA-AP more challenging, which can result in death. Nair and Satheesh have indicated that the incidence of unrecognized DKA-AP accounts for at least 15% of the DKA population and have broadly called for clinicians to screen for DKA-AP by serum amylase, lipase, and triglyceride assessment (1). An analysis of a U.S. population-based survey revealed a higher mortality rate in patients with DKA-AP compared to those with AP (aOR 2.3, P < .001; CI 1.8-3.0). It is noteworthy that this study demonstrated a national incidence of DKA-AP of 1% in the United States between 2003 and 2013, with a cumulative total of 33,356 patients with DKA-AP (2). The presented data indicates that DKA-AP is not an exceedingly uncommon occurrence. The reason for the failure to diagnose DKA-AP, which has resulted in fatalities among patients with DKA or AP, may be attributed to the absence of validated clinical predictive risk factors upon which to base clinical diagnostic decisions. The objective of this study was to examine the risk factors associated with DKA-AP through a retrospective analysis and to evaluate the predictive efficacy of the identified risk factors. This will contribute to the prevention and management of patients with DKA-AP and enhance diagnostic accuracy.

## 2 Methods

This study employed a retrospective design. The clinical data of 45 patients with DKA-AP who were admitted to the emergency department, gastroenterology department, endocrinology department, and critical care medicine department of Yangzhou University Affiliated Hospital between September 2013 and December 2024 were analyzed. To serve as a control group, DKA patients without AP (DKA-nonAP) with an equivalent number of cases during the same period were selected. The control group was matched to the DKA-AP group in terms of age, gender, and duration of diabetes, except for the absence of AP, in order to reduce the influence of confounding factors. The matching process was based on the propensity score matching method, whereby propensity scores were calculated based on demographics, medical history, symptoms, and laboratory parameters. DKA patients with similar scores were selected as the control group. The diagnosis of AP is based on the Guidelines for the Diagnosis and Treatment of Acute Pancreatitis in China (2021). The study was approved by the Institutional Ethics Committee of Yangzhou University Affiliated Hospital (2022-YKL3-06-004).

This study employed a mixed research methodology of retrospective cohort study and systematic review. This mixed research method was similar to that envisaged by Zhu et al(3).

First retrospective cohort study included 90 patients (45 DKA-AP, 45 DKA-nonAP) admitted between 2013–2025. AP diagnosis followed the 2021 Chinese guidelines for acute pancreatitis. The idea of using the Chinese pancreatitis guidelines as a diagnostic criterion for AP is similar to the design of a Chinese DKA-AP study, which reduces study bias in China (4). Data were extracted from electronic health records using a standardized checklist, including demographics, medical history, symptoms, and laboratory parameters.

The literature related to the study of DKA-AP was then reviewed according to the PRISMA guidelines(5). As of February 2025, the terms ‘diabetic ketoacidosis’ and ‘pancreatitis’ were searched using the database PubMed. The search terms ‘diabetic ketoacidosis’ and ‘pancreatitis’ were chosen because they are the two diseases involved in this study and allowed for a precise search of the literature that directly investigated the relationship between DKA and AP. Literature. Also, after the initial search, it was found that these two terms could cover most of the relevant studies. In addition, other synonyms or variants were not selected for the search because it was considered that too many search terms might introduce a large number of irrelevant literature and reduce the efficiency of the search. Further additions can be made if subsequent studies reveal a need to expand the search scope. Only studies examining DAK-AP were included, including clinical randomised controlled studies, cohort studies and observational studies. This was done because we wanted to understand the progress of previous international research on identifying risk factors for DAK-AP. Articles may contain unifactorial analyses, multifactorial analyses, however case reports or case series alone are not sufficient to explore risk factors. Thus case reports and review articles that lacked exploration of risk factors were excluded. We will run backtracking methods applied to the included articles to capture potential literature. The method of performing systematic search and reverse tracking was informed by the method of Zhao et al., Ma et al. and Liang et al., who argued that the heterogeneity of randomised controlled studies is the smallest of all clinical study types and should be considered first, and that the use of reverse tracking method may compensate for the shortcomings in the search formula (6–8). The search was limited to search terms appearing in the title, abstract and keywords to accurately screen the literature studying DKA-AP. Meanwhile, only English literature was included to ensure the quality of literature and language consistency. In addition, the retrieved literature was preliminarily screened to exclude literature that was not directly related to the study of DKA-AP, such as literature that only studied a single disease, DKA or AP, and did not address the association between the two.

Continuous variables were compared using independent t-tests or Mann-Whitney U tests; categorical variables used Pearson’s χ^2^ test. Multivariate logistic regression identified independent risk factors. Receiver operating characteristic curve (ROC) assessed predictive performance. Analyses were performed using SPSS 25.0. Statistical significance was set at bilateral P < 0.05. Post hoc efficacy analyses were conducted to assess the effect of sample size on the study results. We planned to use the G*Power 3.1 software with the principle equation as (**Equation 1**).

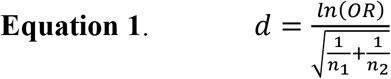

## 3 Results

### 3.1 Baseline Characteristics

A total of 90 patients diagnosed with DKA were included in the study, comprising 45 cases with AP and 45 cases without AP. The DKA-AP group had higher BMI (26.30 vs. 23.20 kg/m^2^, p=0.031), hyperlipidemia prevalence (26.7% vs. 4.4%, p=0.004), and abdominal pain duration (3 vs. 0 days, p<0.001). Significant differences were observed in hematocrit (HCT), red blood cell count (RBC), pH, bicarbonate (HCO_3_^−^), glucose (Glu), total cholesterol (TC), triglycerides (TG), and amylase (AMY) (p<0.05; **Table 1**).

**Table 1.**
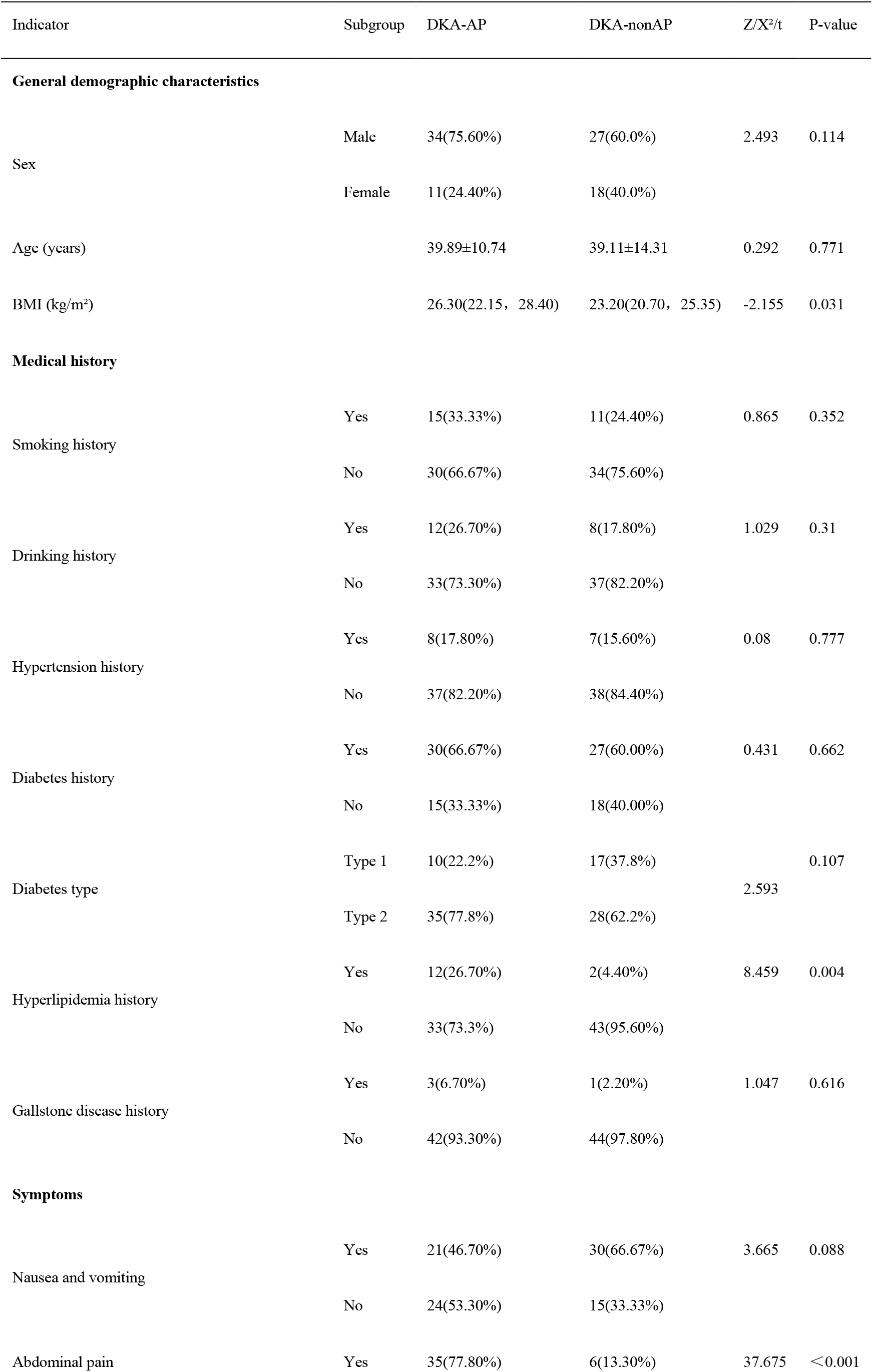
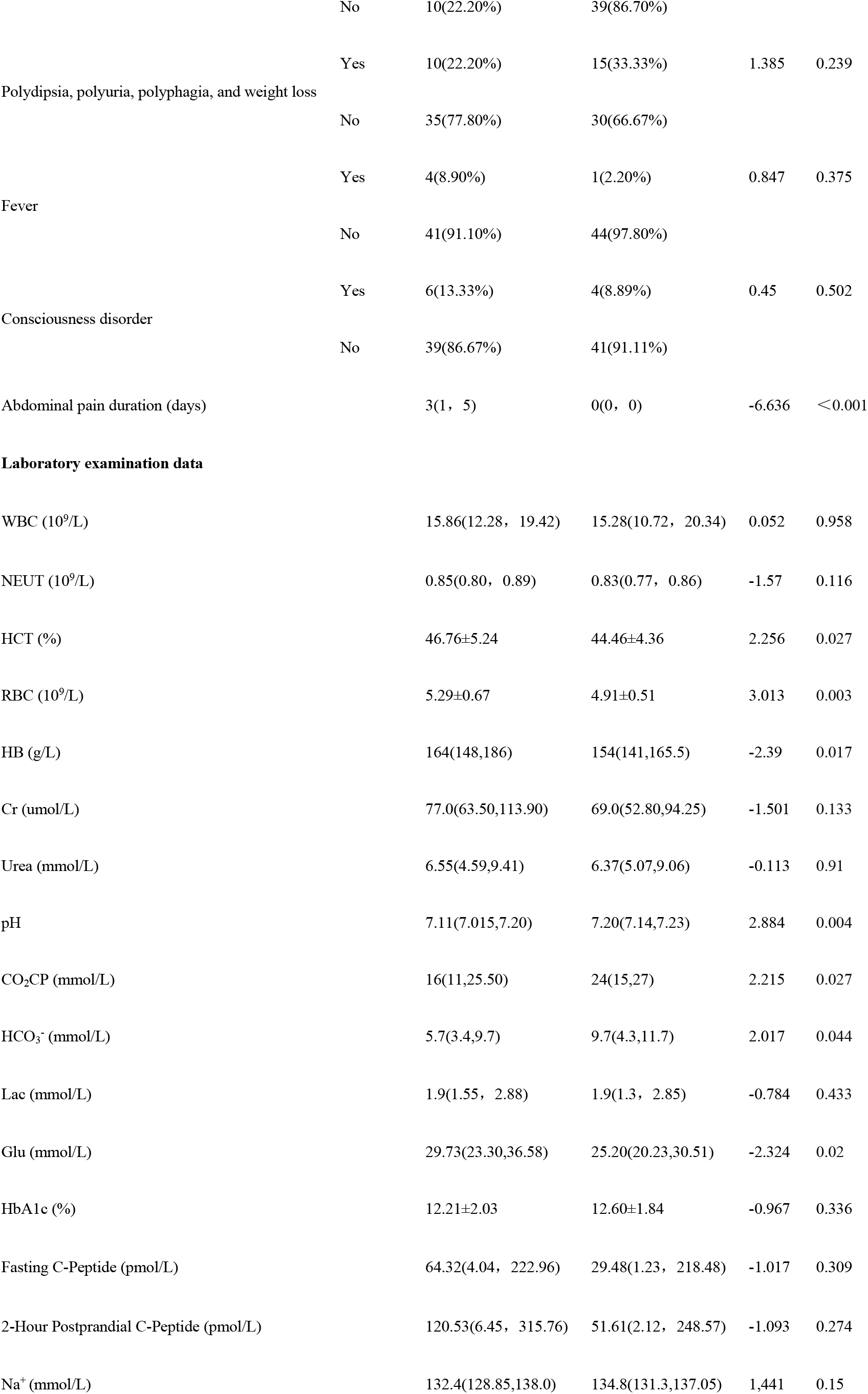
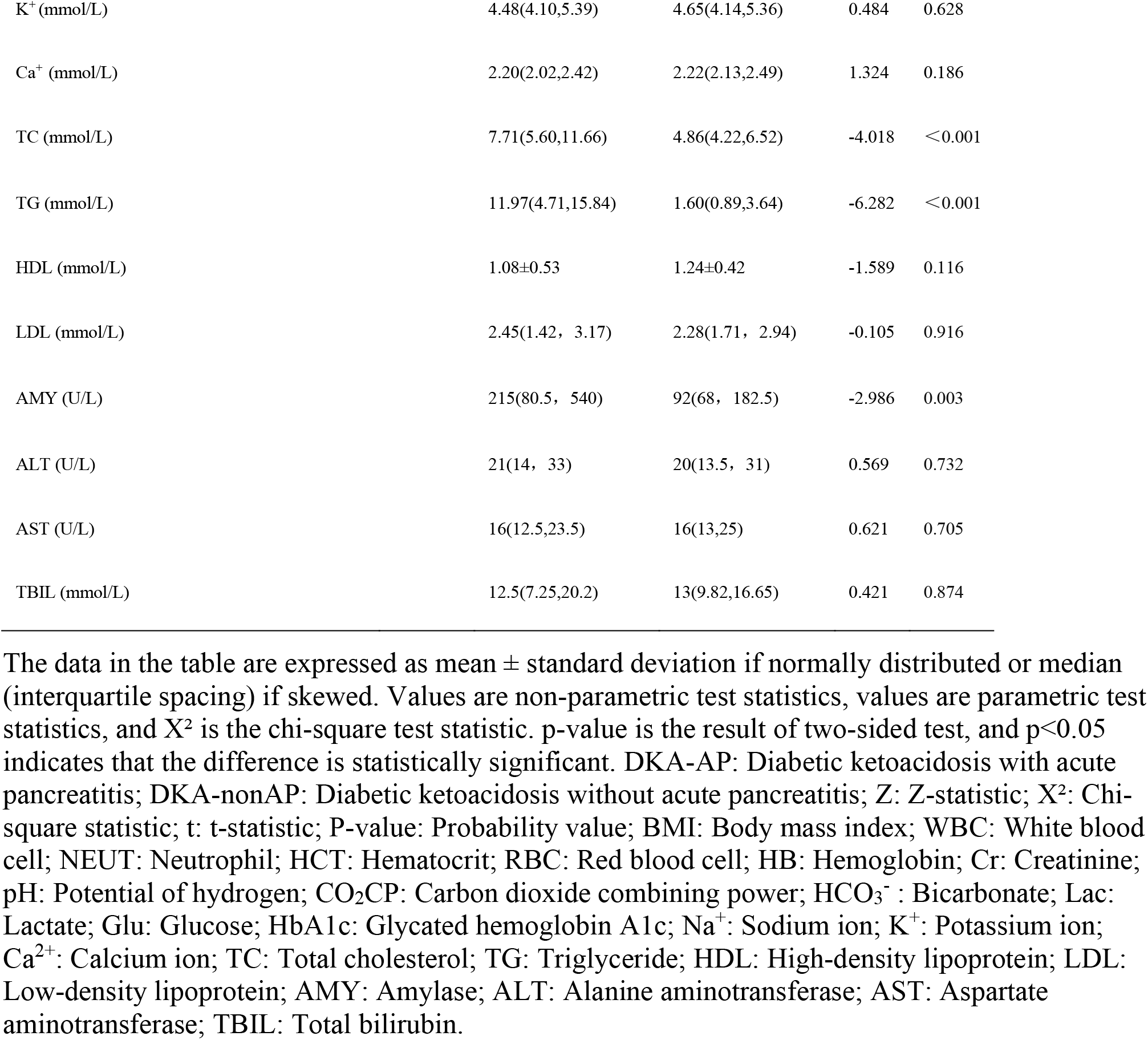
Comparative analysis of the basic conditions of the two groups.

A total of 90 patients diagnosed with DKA were included in the study, comprising 45 cases with AP and 45 cases without AP. The DKA-AP group had higher BMI (26.30 vs. 23.20 kg/m^2^, p=0.031), hyperlipidemia prevalence (26.7% vs. 4.4%, p=0.004), and abdominal pain duration (3 vs. 0 days, p<0.001). Significant differences were observed in hematocrit (HCT), red blood cell count (RBC), pH value(pH), carbon dioxide combining power (CO_2_CP), bicarbonate (HCO_3_^−^), glucose (Glu), total cholesterol (TC), triglyceride (TG), and amylase (AMY) (p<0.05; **Table 1**).

### 3.2 Multivariate analysis

Multivariate analysis was conducted on the indicators with significant differences in the univariate analysis, as illustrated in **Table 2**. The findings revealed that cholesterol, triglyceride levels, and the abdominal pain duration were independent predictors of DKA-AP (**Table 2**). ROC analysis showed high discriminative power (p<0.05; **Table 3**; **Figure 1**).

**Table 2.**
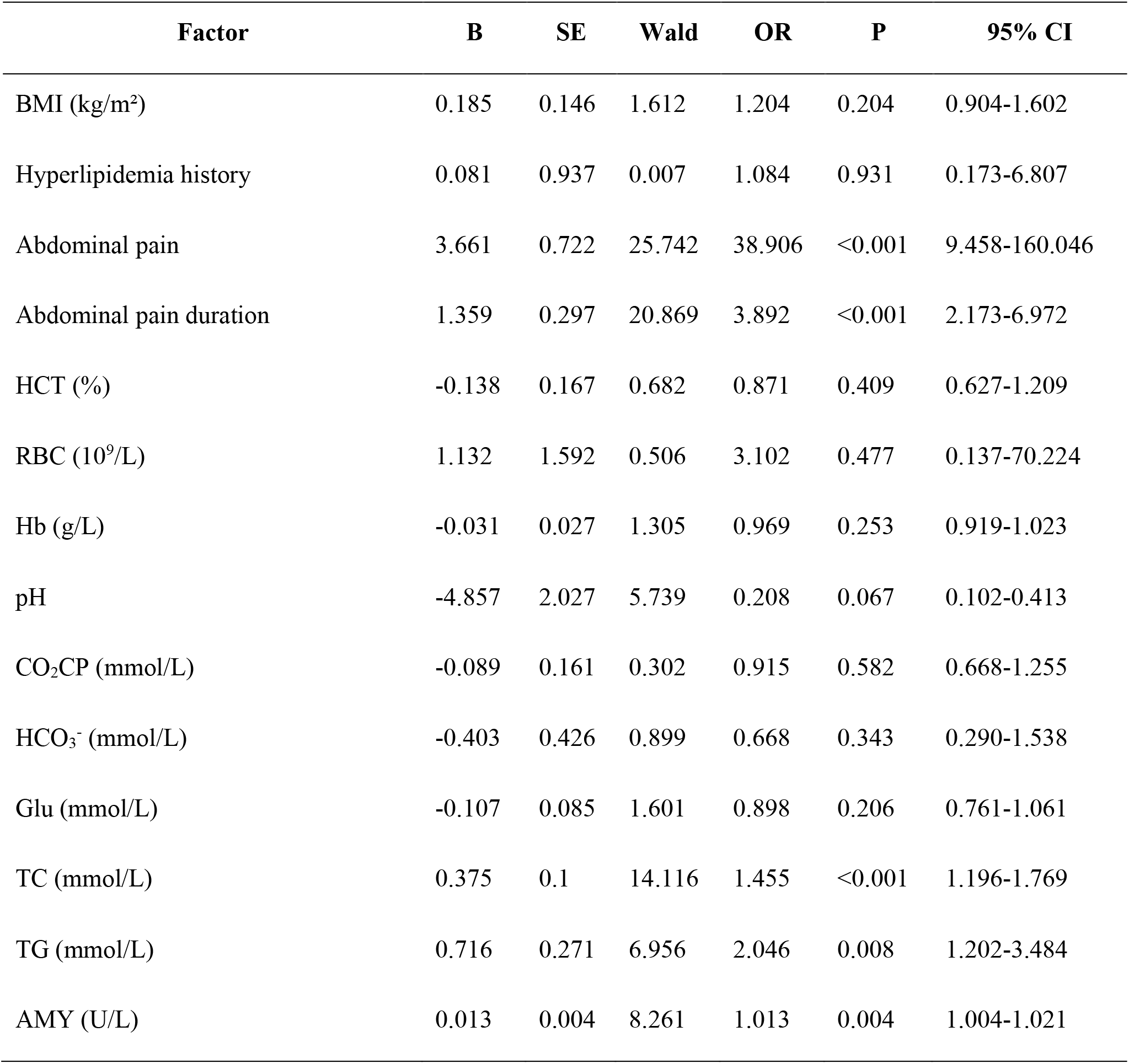
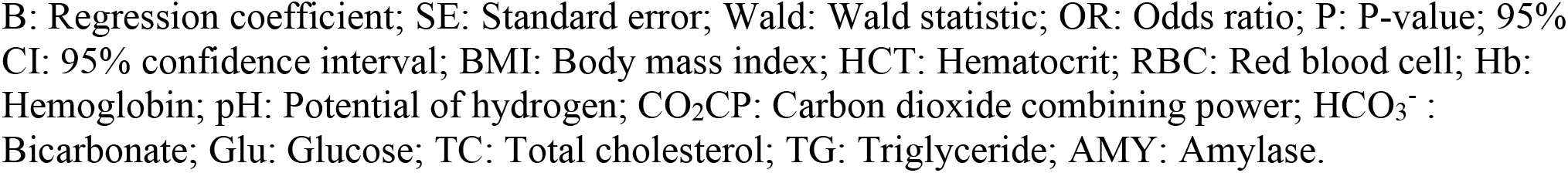
Logistic regression analysis of risk factors associated with DKA-AP.

**Table 3.** Area under the ROC curve for different risk factors.

| Variable | AUC area | Sensitivity (%) | Specificity (%) | Youden's Index | 95% CI | P-value |
| --- | --- | --- | --- | --- | --- | --- |
| TC | 0.751 | 71.1 | 75.6 | 0.467 | 0.648-0.855 | <0.001 |
| TG | 0.884 | 84.4 | 80 | 0.644 | 0.815-0.953 | <0.001 |
| Abdominal pain duration | 0.871 | 77.8 | 86.7 | 0.644 | 0.791-0.951 | <0.001 |
| Combined Diagnosis | 0.933 | 93.3 | 73.3 | 0.667 | 0.881-0.986 | <0.001 |

95% CI: 95% confidence interval; TC: total cholesterol; TG: triglyceride.

**Figure 1.**
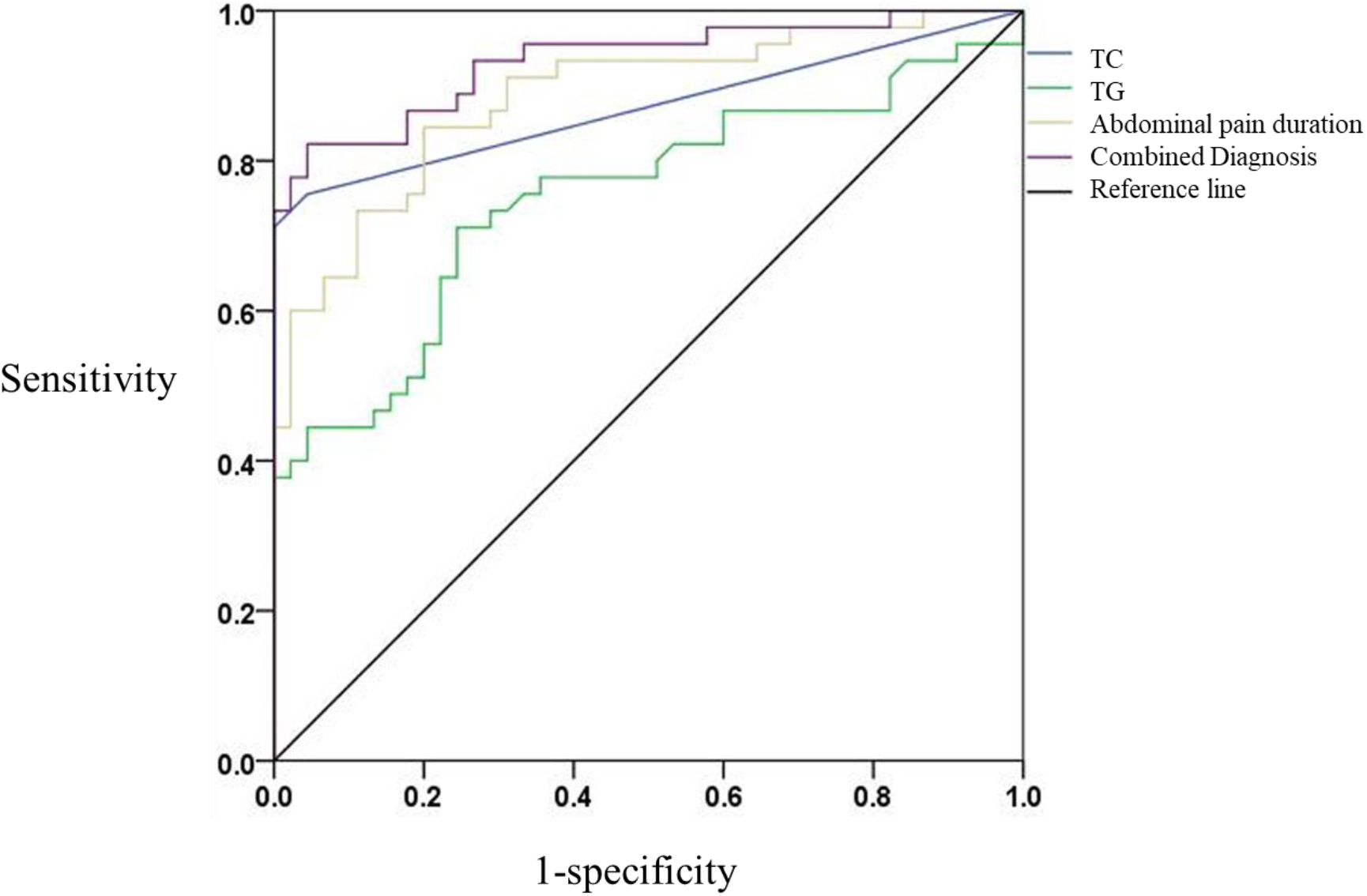
Comparison of ROC curves in subjects with different independent risk factors.

Post hoc efficacy analyses were conducted to assess the effect of sample size on the study results. Using G*Power 3.1 software, an alpha error probability of 0.05 was set, and based on the effect sizes obtained (ORs and corresponding standard deviations for TC, TG, and abdominal pain duration), the statistical efficacy of the study for the main effect was calculated to be 0.54. A statistical efficacy of 0.54 suggests that the probability of the study detecting a true moderate or higher effect with the current sample size is 54%. This means that the study may have some risk of underdiagnosis, i.e., some true moderate effect relationships may not have been detected. For some potential factors associated with DKA-AP, there is a nearly 50-50 chance that they will be missed if they have a moderate effect size. In clinical practice, this may lead to an inadequate assessment of the risk of the condition in some patients, compromising early diagnosis and intervention. Therefore, caution is needed when interpreting the results of the study and highlights the importance of future studies with larger sample sizes.

The horizontal coordinate is the false positive rate (1-specificity) and the vertical coordinate is the true positive rate (sensitivity). The different curves have different meanings. The blue solid line is the TC)predictive ROC curve for DKA-AP; the green solid line is the TG predictive ROC curve for DKA-AP; and the yellow solid line is the abdominal pain duration predictive ROC curve for DKA-AP. The solid purple line represents the ROC curve predicted by the combined diagnosis. The black solid line represents the reference line.

### 3.3 Literature review

After a systematic search in the PubMed database(**Figure 2**), we found 11 clinical studies investigating DKA-AP (1,4,9–17). Cumulatively, 1851 patients were studied, including 3 interventional studies, 1 genetic observational study, and 7 cohort studies exploring risk factors (**Table 4**). Of the 7 cohort studies exploring risk factors, 2 studies analysed risk factors using area under the ROC curve, 2 studies analysed risk factors using a multifactorial approach, and the remaining 3 used a univariate or descriptive method of Analysis.

**Table 4.**
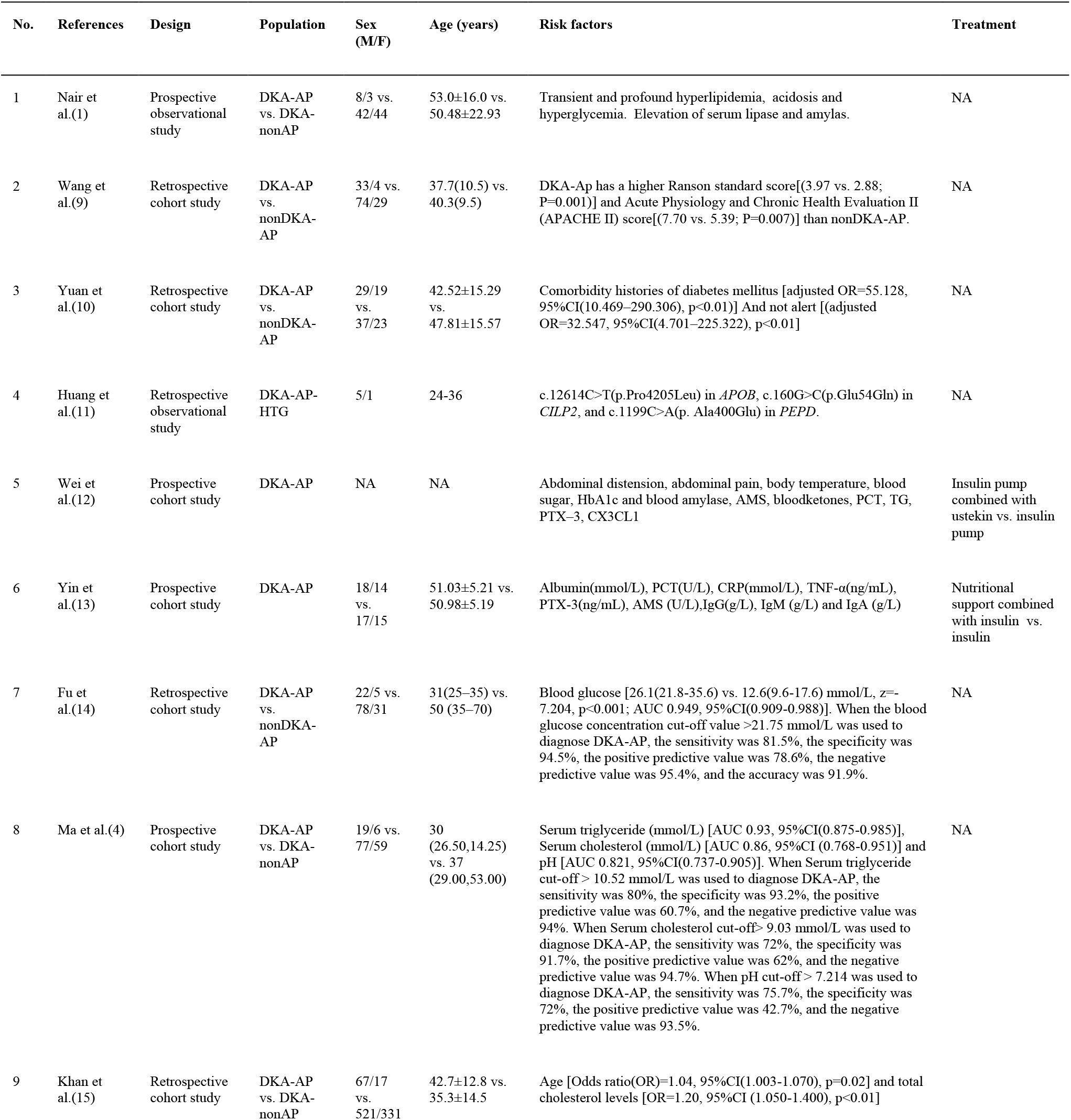
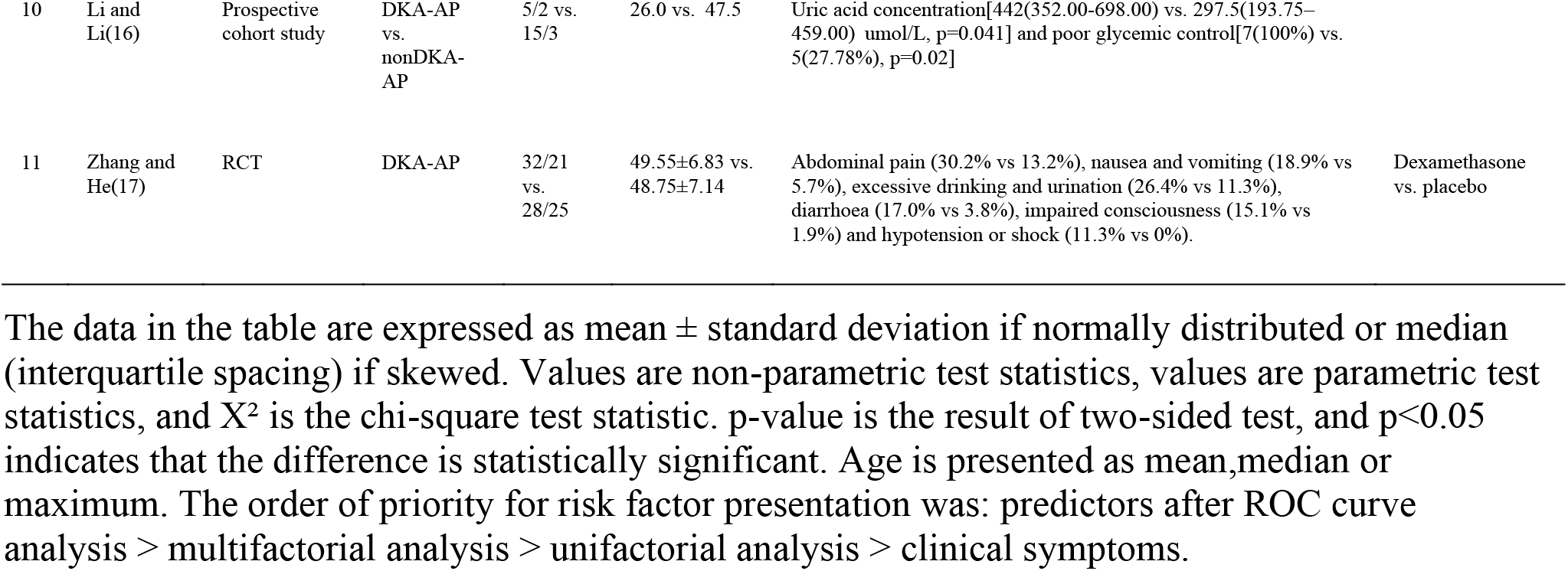
Review of 1851 cases of clinical study of DKA-AP.

**Figure 2.**
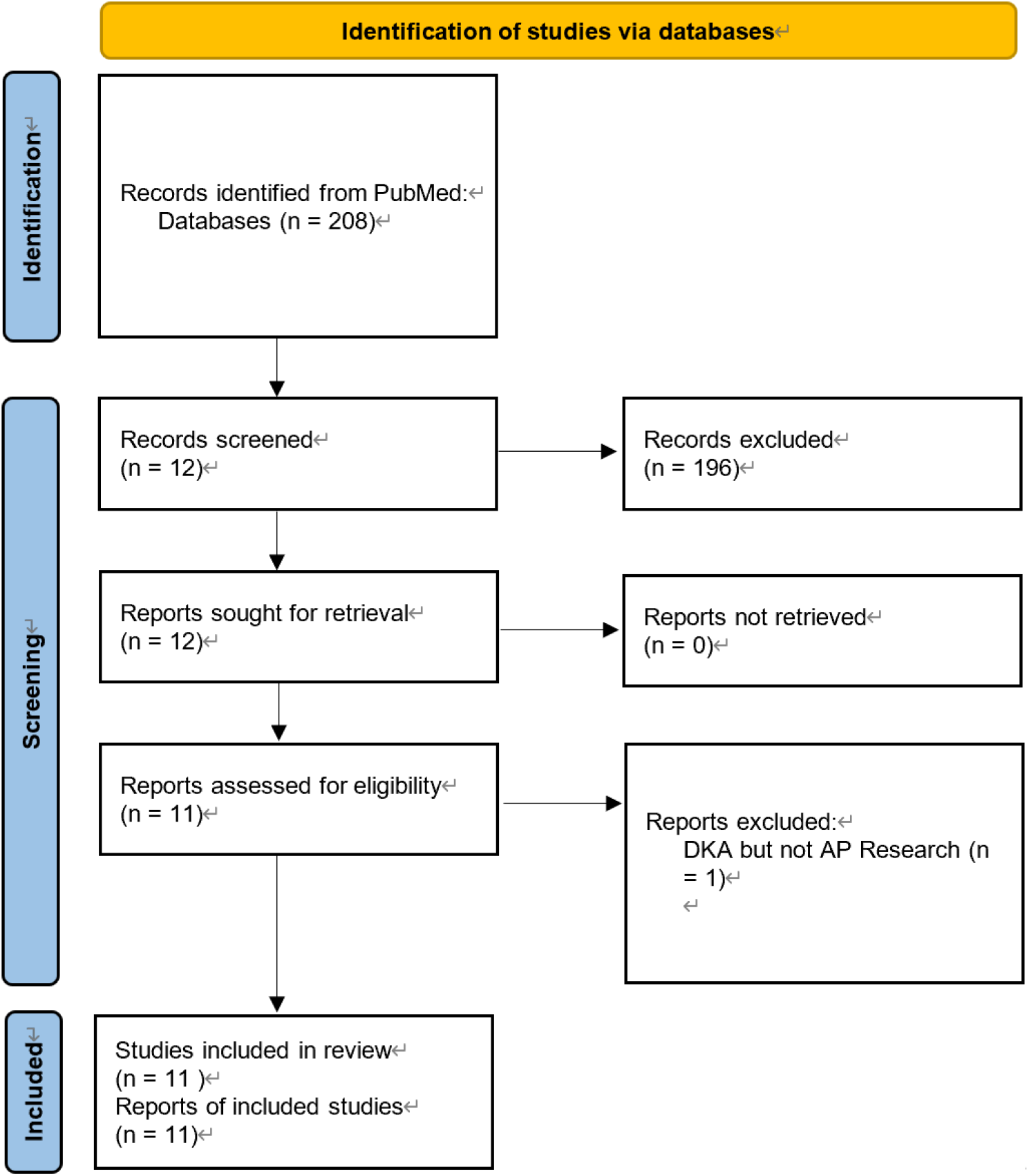
Flowchart of literature selection using the PRISMA guidelines.

From the results, both for DKA and AP, if co-morbidity occurs at the same time, then there seem to be many significant unifactorial difference results in clinical symptoms, clinical assessment scores, laboratory tests and imaging. These differences are not universal and the main reason may be that the population investigated is so small that there is a large single-centre bias. However blood glucose values, total cholesterol levels, age and pH derived from ROC curve analysis can alert the first clinician in the clinic. Khan et al. showed that the younger the age the more likely to be comorbid with AP in patients with DKA (15).

## 4 Discussions

### 4.1 Key results

The aim of this study was to investigate the risk factors for DKA-AP and to assess the predictive efficacy of the identified risk factors. The results of the study showed significant differences between the DKA-AP group and the DKA-nonAP group in a number of indices, with the differences in BMI, history of hyperlipidemia, duration of abdominal pain, and lipid profiles (TC, TG) being particularly prominent. TC, TG levels and duration of abdominal pain were identified as independent predictors of DKA-AP by multifactorial analysis, and ROC curve analysis showed that the combination of these factors had good predictive performance (AUC = 0.933, sensitivity = 93.3%, specificity = 73.3%). In addition, patients with DKA had elevated levels of TG (>11.97 mmol/L), reflecting disturbed lipid metabolism, which may trigger AP through free fatty acid-mediated pancreatic injury. Notably, abdominal pain duration over 3 days became a new predictor. This is in line with the aim of the study and provides an important basis for early clinical recognition of DKA-AP.

### 4.2 Limitations

There are some limitations to this study. Regarding the sample size, only 90 patients were included during the 11-year period, a small sample size, which limits the feasibility of subgroup analyses and may not accurately reflect the situation of patients in different subgroups. For example, it is difficult to carry out in-depth analyses of the differences in risk factors in different gender and age subgroups, which limits the precision of the study results.

On potential confounders, there were unmeasured confounders that could have affected the results, although some were considered in the study design. Patient dietary composition and physical activity were important potential confounders. Chronic high-triglyceride diets may increase the risk of AP development and are also associated with the development and poor control of DKA. Inadequate exercise may lead to metabolic disturbances that promote the development of DKA and AP. As this information was not collected in this study, it could not be adjusted in the analyses, which may have led to biased results. This bias may make the association between risk factors and DKA-AP overestimated or underestimated, interfering with the judgement of the true relationship.

In addition, this study was a single-centre retrospective study with single-centre bias. The single-centre patient population has limitations in terms of geography, ethnicity and medical level, which may not be representative of the wider patient population and affect the generalisability of the study results.

### 4.3 Interpretation

In terms of the achievement of the study objectives, this study successfully targeted TC, TG and abdominal pain duration as the key predictors of DKA-AP. This result is important for achieving the goal of exploring risk factors and provides a key reference for early clinical identification of DKA-AP, which will help clinicians to take timely interventions to improve patients’ prognosis.

However, there are some limitations of the study. Regarding the sample size, this study had a limited sample size, which may lead to instability and bias in the results. Unmeasured factors may have interfered with the judgement of the true relationship between risk factors and DKA-AP. Meanwhile, this study is a single-centre study with limitations in terms of geography, ethnicity and medical environment, which makes it difficult to represent all groups of DKA patients and limits the general applicability of the results.

When compared with similar studies, the results of the present study present similarities and differences. In terms of hypertriglyceridemia as a risk factor for DKA-AP, the findings of several studies are consistent. Nair et al. conducted a prospective study on 100 patients with DKA and found that 11% of them had concomitant AP, and among them, 4 patients with AP had severe hypertriglyceridemia, and their triglyceride levels returned to near normal after remission of the DKA. Nair’s results strongly suggest a strong association between hypertriglyceridemia and DKA-AP(1). Nair’s results strongly demonstrated the strong association between hypertriglyceridemia and DKA-AP(1). Huang et al. conducted a study on 6 patients with DKA and hypertriglyceridemia, and the findings of the present study showed similarities(11). Huang et al. studied six patients with DKA, hypertriglyceridemia, and the AP triad and found that the patients’ triglyceride levels were extremely high, with a mean of 3282.17±2975.43 mg/dL (11), further supporting this view. In addition, Yuan et al. showed a significantly higher incidence of hypertriglyceridemia in AP patients with combined DKA compared to AP patients without DKA. Fu et al. also demonstrated that patients in the DKA group had significantly higher triglyceride levels than those in the non-DKA group (14). Together, these studies confirm the important role of hypertriglyceridemia in the pathogenesis of DKA-AP.

However, there were differences between the present study and other studies regarding the role of factors such as BMI and age. BMI was not identified as an independent risk factor in the present study, whereas Khan et al. showed that BMI was significantly higher in patients with DKA-AP than in patients with DKA-nonAP(15). This may be due to the geographic and ethnic differences in the sample of this study, which was relatively small, resulting in an inadequate ability to detect the effect of BMI. Also, the different ways of measuring and categorising BMI in the study methodology may have contributed to the difference in results.

Regarding age, the present study did not find any significant association between age and DKA-AP, whereas Khan et al. showed that age was a significant risk factor for DKA-AP and that patients with DKA-AP were older(15). However, a study by Fu et al. found that among patients with type 2 diabetes combined with AP, those in the DKA group were younger than those in the non-DKA group(14). This difference may be related to the age range of patients included in each study, type of disease and study design. The age range of patients included in this study may have been narrower, making the effect of age difference on the results not fully apparent; and the different selection criteria for DKA and AP patients in different studies may have led to inconsistencies in the age-DKA-AP relationship. These could interfere with the results of studies of the association of the age factor with DKA-AP.

In addition, other studies have addressed some aspects that were not explored in depth in this study. For example, Wang et al. investigated the effect of DKA on the clinical course and severity scores of hypertriglyceridemia-induced pancreatitis (HP), and found that DKA led to increased acute kidney injury and higher chance of ICU admission in patients with HP (9). Wang’s results are different from the risk factors that were focused on in the present study, but they also suggest that the different aspects of the disease need to be considered comprehensively in the assessment of DKA-AP. The results of Wang’s study are different from the present study but suggest that different aspects of the disease need to be considered together when assessing DKA-AP. Other studies have focused on the effect of treatment modalities on patients with DKA-AP. Wei et al. found that insulin pumps combined with ustekin therapy reduced PCT, TG, and other markers in patients (12). Yin et al. demonstrated that nutritional support combined with insulin therapy improved serum protein levels and reduced inflammatory response in patients (13). These studies provide a reference for the clinical management of DKA-AP from a therapeutic point of view, and also show that when interpreting the results of the risk factors in this study, it is necessary to make a comprehensive judgement in combination with the impact of different treatment modalities on the disease.

In addition, multiple analyses were conducted in the study, and although methods such as multivariate logistic regression analyses were used to identify independent risk factors and ROC curves were used to assess the predictive performance, there may be some limitations and uncertainties in the different analytical methods. When combining the results of multiple analyses, there may also be contradictory or difficult-to-interpret situations.

Although this study has achieved some results in exploring the risk factors of DKA-AP, clinicians should be cautious in applying the results of this study due to the limitations of the study, the complexity of multiple analyses, and the differences with similar studies. The results cannot be directly generalised to all DKA patients, but should be used in conjunction with the individual patient’s condition, the characteristics of the region, and other clinical evidence to formulate a more accurate and effective diagnosis and treatment plan. Future studies need to expand the sample size, conduct multicentre studies, and include more potential confounding factors to further validate and improve the findings of this study.

## 5 Conclusions

Elevated TC, TG, and prolonged abdominal pain duration are critical predictors of DKA-AP. These factors enhance diagnostic accuracy and guide targeted management, reducing morbidity in high-risk populations. Future multi-center studies with larger cohorts are warranted to validate these predictors and explore additional risk factors, such as genetic or lifestyle-related variables. At the same time, a prospective study design can be adopted to track the changes of patients’ conditions in depth and establish a better risk prediction model to provide a stronger basis for early clinical diagnosis and intervention.

## Data Availability

All data produced in the present work are contained in the manuscript.

## 7 Author Contributions

YB: Writing – review & editing, Conceptualization, Methodology, Formal Analysis, Project administration; MC: Writing – review & editing, Conceptualization, Methodology; YF: Writing – original draft, Conceptualization, Methodology; HZ: Writing – review & editing.

## Acknowledgments

We thank the emergency department, gastroenterology department, endocrinology department, and critical care medicine department of Yangzhou University Affiliated Hospital for supporting our clinical research. We also thank Northwest Minzu University, Lanzhou, China, for providing evidence-based medicine assistance. The study was approved by the Institutional Ethics Committee of Yangzhou University Affiliated Hospital (2022-YKL3-06-004).

